# Medicaid Expansion and Survival Outcomes among Men with Prostate Cancer

**DOI:** 10.1101/2024.10.23.24315994

**Authors:** Oluwasegun Akinyemi, Mojisola Fasokun, Eric Hercules, Seun Ikugbayigbe, Eunice Odusanya, Nadia Hackett, Oluebubechukwu Eze, Lerone Ainsworth, Kakra Hughes, Edward Cornwell, Pamela Coleman

## Abstract

**INTRODUCTION:** Prostate cancer stands as one of the most diagnosed malignancies among men worldwide. With the recent expansion of Medicaid under the Affordable Care Act (ACA), millions more Americans now have health insurance coverage, potentially influencing healthcare access and subsequent outcomes for various illnesses, including prostate cancer. Yet, the direct correlation between Medicaid expansion and cancer-specific survival, particularly for early-stage prostate cancer, remains an area warranting comprehensive exploration.

**OBJECTIVE:** This study aims to determine the impact of the implementation of Medicaid expansion on Survival outcomes among men with prostate cancer.

**METHODS:** We utilized data from the SEER registry to determine the causal impact of the implementation of the ACA on outcomes among men with prostate cancer. The study covered the years 2003-2021, divided into pre-ACA (2003-2009) and post-ACA (2015-2021) periods, with a 1-year washout (2014-2015) since Medicaid expansion was implemented in 2014 in Kentucky. Using a Difference-in-Differences approach, we compared survival among men with prostate cancers from Kentucky to Georgia. We adjusted for patient demographics, income, metropolitan status, disease stage, and treatment modalities.

**RESULTS:** We analyzed a cohort of 68,222 men with prostate cancer during the study period. Of these, 37,810 (55.4%) were diagnosed in the pre-ACA period, with 70.8% from Georgia and 29.2% from Kentucky. The remaining 30,412 (44.6%) were diagnosed in the post-ACA period, with 72.3% from Georgia and 27.7% from Kentucky. Medicaid expansion in Kentucky was associated with a 16.8% reduction in hazard of death (HD), indicating improved overall survival among low-income individuals. This trend was consistent across different racial/ethnic groups. Specifically, Non-Hispanic white men experienced a 16.2% reduction (DID = −0.162, 95% CI: − 0.315 to −0.008), Non-Hispanic Black men had a 17.9% reduction (DID = −0.179, 95% CI: −0.348 to −0.009), and Hispanic men saw a 15.9% reduction (DID = −0.159, 95% CI: −0.313 to − 0.005) in HD among low-income individuals.

**CONCLUSION:** Medicaid Expansion was associated with a substantive improvement in overall survival among men with prostate cancers in Kentucky compared to non-expansion Georgia.

## INTRODUCTION

Outside of skin cancer, prostate cancer is the most common cancer in men in the United States (U.S.) [1], with approximately 299,010 new cases of prostate cancer expected in 2024, along with about 35,250 deaths from the disease [2]. Since 2014, the incidence rate has sharply increased 3% per year overall and 5% per year for advanced-stage prostate cancer [2]. For men diagnosed with prostate cancer, the treatment can range from observation, curative surgery (prostatectomy), and targeted radiation therapy [3], however, what is not often discussed are the factors that determine the treatment of choice [3,4]. Financial toxicity is a term that is used to describe the financial consequences and the potential stress incurred by a disease diagnosis and treatment [5]. In 2020 alone, there was an estimated $22.3 billion spent on prostate cancer care, with the majority of that being made up by direct medical expenses [6]. However, the economic impact goes far beyond medical bills. Prostate cancer leads to very significant lost earnings as a result of premature mortality and the inability of patients to work during their treatment and recovery process [7].

Prostate cancer is a very complex disease whose pathogenesis involves both genetic and environmental factors and is mainly driven by alterations in androgen signaling [8]. There are a multitude of risk factors that contribute to the development of prostate cancer, including age, family history, and race [9]. African American men are at a higher risk of developing prostate cancer and usually tend to present with a more advanced disease at diagnosis compared to their counterparts [10]. It has also been shown that social determinants of health (SDOH) have a significant impact on prostate cancer outcomes(11). Factors such as socioeconomic status, access to healthcare, and living conditions not only influence the state at which prostate cancer is diagnosed, but also overall survival rates [12]. Prior data suggests that incidence rates are associated with socioeconomic status [10]. Additionally, lower socioeconomic status is associated with poorer survival rates [10]. In order to reduce disparities in prostate cancer care and improve survival, it is crucial to address the SDOH that may be impacting the outcomes of disease.

The goal of Medicaid, initially enacted in 1965, was to allow States to receive federal funding to provide health insurance to persons with limited income [13]. This legislation was later expanded and termed The Affordable Care Act (ACA), passed in 2010 and fully implemented in 2014 [14]. This legislation addressed Americans without health insurance facing systemic health inequalities [14]. This act is the most significant expansion of coverage in the US healthcare system following the creation of Medicare and Medicaid in 1965 [15]. Before the ACA, there were limitations to Medicaid eligibility on the state level, which covered only the poorest in specific categories (i.e., disabled people, pregnant women, and children) [15]. In contrast, with the ACA, citizens with income at or below 138% of the federal poverty line qualify for aid [14]. Due to ACA, the number of uninsured individuals declined significantly from 2013 to 2022, from approximately 45.2 million to 26.4 million [5]. The overall implementation of the Affordable Care Act during 2013-2022 has led to an overall 45% increase in Americans with health insurance coverage [16].

When looking specifically at the effect of the ACA on patients newly diagnosed with breast, colorectal, and lung cancer from 2012 to 2015 there has been an increase in earlier stage of diagnosis and lower hazard of mortality among patients [17]. A study done written by Eugia et al assessed the effect of Medicaid expansion in states with coverage in patients with bladder, colorectal, esophageal, lung, bladder and gastric cancers and found that there was a increase in number patients who had access to healthcare and noted a positive effect on access and utilization of oncologic care [18]. We understand the significance of the implementation of the affordable care act on the improved access of care utilization, however direct correlation between Medicaid expansion and cancer-specific survival, particularly for early-stage prostate cancer, remains an area warranting comprehensive exploration. This study aims to determine the impact of the implementation of the Affordable Care Act on cancer-specific survival among men with early-stage prostate malignancies.

## METHODOLOGY

### Data Source and Study Population

This retrospective cohort study utilized data from the Surveillance, Epidemiology, and End Results (SEER) database, focusing on men diagnosed with prostate cancer between 2000 and 2021[19]. SEER collects cancer incidence and survival data from population-based cancer registries covering approximately 48% of the U.S. population as of 2021 [20]. The study utilized the “Incidence - SEER 18 Registries, Nov 2021 Sub (2000–2021)” database [19]. Inclusion criteria included: (1) men diagnosed with prostate malignancies identified using International Classification of Diseases for Oncology, 3rd edition (ICD-O-3) codes; (2) patients with complete clinicopathological information and survival data; (3) age at diagnosis between 18 - 64yrs; and (4) patients actively followed up. Exclusion criteria were: (1) patients diagnosed through autopsy or death certificate only, or with clinical diagnoses only; (2) patients with missing data on race/ethnicity, household median income, state identification, or years of follow-up. This study involved publicly available data and did not include any identifiable patient information, so institutional review board approval was not required. Informed consent was also not necessary due to the retrospective nature of the study.

### Georgia vs. Kentucky

Georgia (control state) and Kentucky (treated state) were selected for the study due to their contrasting approaches to healthcare policy, specifically regarding the implementation of the Affordable Care Act (ACA). Kentucky expanded Medicaid under the ACA in 2014, leading to a significant increase in Medicaid/CHIP enrollment and a reduction in the uninsured rate. In contrast, Georgia did not expand Medicaid, resulting in higher uninsured rates and more restrictive access to healthcare services. The differences in healthcare policy between the two states provide a unique opportunity to assess the impact of the ACA on prostate cancer outcomes, particularly cancer-specific survival and overall survival.

### Primary Outcome of Interest

The primary outcomes of interest were cancer-specific survival (CSS) and overall survival (OS). CSS was defined as the time from diagnosis to death due to prostate cancer, while OS referred to the time from diagnosis to death from any cause. Both outcomes were assessed over two periods: pre-ACA (2003-2009) and post-ACA (2015-2021).

### Independent Variables of Interest

The primary independent variables were the implementation of the ACA and Medicaid expansion, categorized into two time periods: pre-ACA (2003-2009) and post-ACA (2015-2021) with a 1-year washout period of January 2014- December 2014 to allow for the policy to become fully operational. Kentucky’s Medicaid expansion served as the treatment, while Georgia, which did not expand Medicaid, served as the control. The interaction between state and time period (State X ACA) was used to evaluate the differential impact of the ACA on prostate cancer outcomes between the two states.

### Covariates

Covariates included demographic factors such as age at diagnosis (continuous variable), race/ethnicity (categorized as Non-Hispanic White people, Non-Hispanic Black people, Hispanic people, Non-Hispanic Asian/Pacific Islander people, and Native American people), and marital status (categorized as married, widowed, divorced, separated, or unknown). Socioeconomic status was assessed using household median income (>$64,000 and ≤$64,000). Tumor characteristics, including stage at diagnosis (localized, regional, distant), and treatment modalities (prostatectomy, chemotherapy, radiation) were also included as covariates.

### Theoretical Model: Andersen Behavioral Model

This study uses the Andersen Behavioral Model of Health Services [21, 22] Use to explore the impact of the ACA and Medicaid expansion on prostate cancer mortality, comparing Kentucky (which implemented Medicaid expansion in 2014) and Georgia (which has not). The model categorizes factors into predisposing, enabling, and need factors:

Predisposing Factors: These include demographics such as age (18-45, 45-64 years), race/ethnicity (White, Black, Hispanic, etc.), and marital status (single, married, etc.), which influence healthcare-seeking behavior.

Enabling Factors: These are resources that facilitate access to care, including income (<64K, ≥64K), metropolitan status (rural, small, medium, large metropolitan), and the state of residence (Louisiana vs. Georgia), reflecting the impact of Medicaid expansion.

Need Factors: These include clinical variables such as cancer grade (Stage I, Stage II, Stage III and Stage IV), and the receipt of treatments like surgery, chemotherapy, and radiotherapy, all of which influence prostate cancer outcomes.

### Difference-in-Differences (DID) Specification

The present study used a DID model to estimate the impact of Medicaid expansion on prostate cancer outcomes; overall survival, overall deaths, and disease stage at presentation by comparing Kentucky (treatment group, which implemented Medicaid expansion) to Georgia (control group, which did not) during the pre-ACA and post-ACA periods. The variable ACA was set to 1 for the post-Medicaid expansion period (2015–2020) and 0 for the pre-expansion period (2003-2009). The variable State was defined as 1 if the observation was from Kentucky (expansion state) and 0 if from Georgia (non-expansion state).

The DID model is specified as:

*y*=*X*β+β_1_⋅ACA+β_2_⋅State + β_12_⋅ (ACA X State) +u [23]

*y* represents the breast cancer outcome of interest (e.g., survival, overall mortality, or disease stage at presentation).

*X* includes covariates such as age, race, marital status, and treatment modalities.

β_1_ captures the difference in outcomes between the pre- and post-ACA periods across both states.

β_2_ captures the baseline difference between Louisiana and Georgia.

β_12_ represents the effect of Medicaid expansion on breast cancer outcomes.

The interaction term ACA × State (i.e., β_12_) estimates the difference in the change in outcomes between Louisiana (the expansion state) and Georgia (the non-expansion state) from the pre- to the post-ACA period. This term provides the key estimate of the impact of Medicaid expansion.

### Statistical Analysis

Descriptive statistics summarized the characteristics of the study population. Chi-square tests were used for categorical variables, and appropriate regression models, such as the Cox proportional hazards model, were applied to time-to-event data. Margins plots were generated to visualize the adjusted probabilities of each outcome across states and time periods. Subsequently, the “lincom” command was used to calculate the DID and generate the standard error, p-value, and confidence intervals. Statistical significance was determined using two-tailed tests with an alpha level of 0.05. All analyses were conducted using STATA 16 statistical software.

## RESULT

### Baseline Study Characteristics

In Table 1, the baseline characteristics of individuals with prostate cancer in Georgia and Kentucky were compared across two time periods, pre-ACA (2003-2009) and post-ACA (2015-2021). In the pre-ACA period, the mean age of diagnosis was similar between the two states, with Georgia at 57.3 ± 5.2 years and Kentucky at 57.6± 5.0 years. Post-ACA, the mean age slightly increased in both states, with Georgia at 58.1± 4.8 years and Kentucky at 58.4 ± 4.7 years. In terms of race and ethnicity, Georgia had a significantly higher proportion of Non-Hispanic Black people compared to Kentucky in both periods. Pre-ACA, 37.0% of patients in Georgia were Non-Hispanic Black people, compared to only 10.5% in Kentucky. Post-ACA, the proportion of Non-Hispanic Black people in Georgia further increased to 47.7%, while in Kentucky, it remained relatively low at 14.2%.

**Table 1:** Demographic, Clinical, and Treatment Characteristics of Individuals with Prostate Cancer in Georgia and Kentucky Before and After Medicaid Expansion in Kentucky in 2014.

| Variables | Pre-ACA (2003-2009)<br>(N=37,810) |  |  | Post-ACA (2015-2021)<br>(N=30,412) |  |  |
| --- | --- | --- | --- | --- | --- | --- |
|  | Georgia<br>(N=26,772) | Kentucky<br>(N=11,038) | P-value | Georgia<br>(N=48,910) | Kentucky<br>(N=19,312) | P-value |
| Age (years; Mean ± SD) | 57.3 ± 5.2 | 57.6 ± 5.0 | <0.01 | 58.1 ± 4.8 | 58.4 ± 4.7 | <0.01 |
| Age |  |  |  |  |  |  |
| 18-44yrs. | 567 (2.1%) | 163 (1.5%) | <0.01 | 251 (1.1%) | 72 (0.9%) | 0.046 |
| 45-64yrs. | 26,205 (97.9%) | 10,875 (98.5%) |  | 21,887 (98.9%) | 8,202 (99.1%) |  |
| Race/Ethnicity |  |  | <0.01 |  |  | <0.01 |
| White Person | 16,158 (60.4%) | 9,698 (87.9%) |  | 10,366 (46.8%) | 6,679 (80.7%) |  |
| Black Person | 9,895 (37.0%) | 1,163 (10.5%) |  | 10,566 (47.7%) | 1,171 (14.2%) |  |
| Hispanic Person | 453 (1.7%) | 57 (0.5%) |  | 740 (3.3%) | 80 (1.0%) |  |
| Other | 266 (1.0%) | 120 (1.1%) |  | 466 (2.1%) | 344 (4.2%) |  |
| Income |  |  | <0.001 |  |  | <0.001 |
| <65K | 12,679 (47.4%) | 9,432 (85.5%) |  | 9,739 (44.0%) | 7,008 (84.7%) |  |
| ≥65K | 14,093 (52.6%) | 1,606 (14.6%) | | 12,399 (56.0%) | 1,266 (15.3%) | |
| Marital Status |  |  | <0.001 |  |  | <0.01 |
| Single | 2,944 (11.0%) | 754 (6.8%) |  | 3,465 (15.7%) | 968 (11.8%) |  |
| Married | 18,772 (70.1%) | 7,358 (66.7%) |  | 12,862 (58.3%) | 4,883 (59.3%) |  |
| Widowed | 507 (1.9%) | 196 (1.8%) |  | 331 (1.5%) | 162 (2.0%) |  |
| Divorced | 2,277 (8.5%) | 997 (9.0%) |  | 1,566 (7.1%) | 845 (10.3%) |  |
| Separated | 273 (1.0%) | 56 (0.5%) |  | 197 (0.9%) | 46 (0.6%) |  |
| Unknown | 1,988 (7.5%) | 1,677 (15.2%) |  | 3,649 (16.5%) | 1,334 (16.2%) |  |
| Metropolitan Status |  |  | <0.001 |  |  | <0.001 |
| Rural areas not adjacent to metropolitan areas | 1,081 (4.0%) | 2,464 (22.3%) |  | 930 (4.2%) | 1,900 (23.0%) |  |
| Rural areas adjacent to metropolitan areas | 3,648 (13.6%) | 1,809 (16.4%) |  | 2,437 (11.0%) | 1,273 (15.4%) |  |
| Small metropolitan areas | 4,037 (15.1%) | 925 (8.4%) |  | 3,020 (13.6%) | 793 (9.6%) |  |
| Medium metropolitan areas | 3,005 (11.2%) | 1,704 (15.4%) |  | 2,459 (11.1%) | 1,262 (15.3%) |  |
| Large metropolitan areas | 15,001 (56.0%) | 4,136 (37.5%) |  | 13,292 (60.0%) | 3,046 (36.8%) |  |
| Stage at presentation |  |  | <0.01 |  |  | <0.01 |
| Localized | 15,383 (85.8%) | 5,731 (80.5%) |  | 6,855 (77.2%) | 2,395 (68.5%) |  |
| Regional | 1,802 (10.1%) | 1,081 (15.2%) |  | 1,237 (13.9%) | 557 (15.9%) |  |
| Distant | 480 (2.7%) | 196 (2.8%) |  | 463 (5.2%) | 160 (4.6%) |  |
| Unknown | 271 (1.5%) | 109 (1.5%) |  | 328 (3.7%) | 384 (11.0%) |  |
| Surgery, n (%) |  |  | <0.001 |  |  | <0.01 |
| Yes | 14,217 (54.0%) | 3,635 (35.7%) |  | 13,867 (63.0%) | 4,088 (54.8%) |  |
| None/Unknown | 12,135 (46.1%) | 6,551 (64.3%) |  | 8,143 (37.0%) | 3,369 (45.2%) |  |
| Chemotherapy, n (%) |  |  | 0.345 |  |  | 0.106 |
| Yes | 191 (0.7%) | 69 (0.6%) |  | 445 (2.0%) | 191 (2.3%) |  |
| None/Unknown | 26,581 (99.2%) | 10,969 (99.4%) |  | 21,693 (98.0%) | 8,083 (97.7%) |  |
| Radiotherapy, n (%) |  |  | <0.01 |  |  | <0.01 |
| Yes | 7,994 (34.2%) | 1,908 (19.5%) |  | 5,210 (25.5%) | 2,197 (27.6%) |  |
| None/Unknown | 15,390 (65.8%) | 7,898 (80.5%) |  | 15,242 (74.5%) | 5,766 (72.4%) |  |

In terms of tumor stage at diagnosis, the proportion of localized cases decreased significantly in both states post-ACA. In Georgia, localized cases dropped from 85.8% pre-ACA to 77.2% post-ACA, while in Kentucky, they decreased from 80.5% to 68.5%. Additionally, the incidence of distant-stage prostate cancer increased in both states post-ACA, rising from 2.7% to 5.2% in Georgia and from 2.8% to 4.6% in Kentucky. Metropolitan status also revealed significant differences, with a higher proportion of patients in Kentucky residing in rural areas compared to Georgia during both periods. For example, in the post-ACA period, 23.0% of patients in Kentucky lived in rural areas not adjacent to metropolitan areas, compared to only 4.2% in Georgia.

### Factor associated with CSS

Table 2 reveals factors associated with CSS among individuals with prostate cancer; comparing Georgia and Kentucky across pre-ACA and post-ACA periods. The post-2015 period was associated with improved survival outcomes, with a hazard ratio (HR) of 0.78 (95% CI: 0.68– 0.89), indicating a significant reduction in the risk of cancer-specific mortality. Additionally, the interaction between states and the post-Medicaid expansion period revealed that individuals in Kentucky had a substantial reduction in mortality post-expansion period (HR = 0.71, 95% CI: 0.56–0.90), suggesting a beneficial effect of the policy in reducing cancer-specific mortality in this state.

**Table 2:** Cox Proportional Hazards Model Assessing Factors Associated with Cancer-Specific Survival among Individuals with Prostate Cancer.

| Variables | Haz. Ratio | Std. Err. | z | P>z | [95% Conf. Interval] |  |
| --- | --- | --- | --- | --- | --- | --- |
| State |  |  |  |  |  |  |
| Kentucky (ref. Georgia) | 1.181 | 0.039 | 5.050 | 0.000 | 1.107 | 1.259 |
| ACA |  |  |  |  |  |  |
| Post-ACA (ref. Pre-ACA) | 0.901 | 0.042 | -2.260 | 0.024 | 0.823 | 0.986 |
| State x ACA |  |  |  |  |  |  |
| Kentucky x Post-ACA | 0.868 | 0.067 | -1.850 | 0.065 | 0.747 | 1.009 |
| Age |  |  |  |  |  |  |
| 18-44yrs. | Reference |  |  |  |  |  |
| 45-64yrs. | 2.175 | 0.283 | 5.980 | 0.000 | 1.686 | 2.806 |
| Race/Ethnicity |  |  |  |  |  |  |
| White Person | Reference |  |  |  |  |  |
| Black Person | 1.103 | 0.031 | 3.480 | 0.001 | 1.044 | 1.166 |
| Hispanics | 0.983 | 0.103 | -0.170 | 0.867 | 0.801 | 1.206 |
| Other | 0.741 | 0.162 | -1.370 | 0.171 | 0.482 | 1.139 |
| Household Median Income |  |  |  |  |  |  |
| Income ≥ 64K (ref. < 64K) | 0.878 | 0.030 | -3.840 | 0.000 | 0.822 | 0.938 |
| Marital Status |  |  |  |  |  |  |
| Single | Reference |  |  |  |  |  |
| Married | 0.649 | 0.023 | -11.960 | 0.000 | 0.605 | 0.697 |
| Widowed | 1.126 | 0.091 | 1.470 | 0.141 | 0.961 | 1.319 |
| Divorced | 0.994 | 0.046 | -0.130 | 0.895 | 0.908 | 1.088 |
| Separated | 1.060 | 0.112 | 0.550 | 0.581 | 0.861 | 1.305 |
| Unknown | 0.751 | 0.040 | -5.430 | 0.000 | 0.678 | 0.833 |
| Metropolitan Status |  |  |  |  |  |  |
| Rural areas not adjacent to metropolitans | Reference |  |  |  |  |  |
| Rural areas adjacent to metropolitans | 0.964 | 0.045 | -0.770 | 0.441 | 0.880 | 1.058 |
| Small metropolitan areas | 0.819 | 0.041 | -3.990 | 0.000 | 0.743 | 0.904 |
| Medium metropolitan areas | 0.776 | 0.039 | -5.060 | 0.000 | 0.703 | 0.856 |
| Large metropolitan areas | 0.713 | 0.033 | -7.340 | 0.000 | 0.652 | 0.781 |
| Grade |  |  |  |  |  |  |
| Localized | Reference |  |  |  |  |  |
| Regional | 1.133 | 0.099 | 1.430 | 0.154 | 0.954 | 1.345 |
| Distant | 3.482 | 0.387 | 11.230 | 0.000 | 2.801 | 4.329 |
| Stage at Presentation |  |  |  |  |  |  |
| Stage I | Reference |  |  |  |  |  |
| Stage II | 1.449 | 0.168 | 3.200 | 0.001 | 1.155 | 1.819 |
| Stage III | 2.135 | 0.318 | 5.100 | 0.000 | 1.595 | 2.858 |
| Stage IV | 4.047 | 0.625 | 9.060 | 0.000 | 2.991 | 5.477 |
| Prostatectomy | 0.473 | 0.017 | -21.050 | 0.000 | 0.441 | 0.507 |
| Chemotherapy | 1.421 | 0.097 | 5.130 | 0.000 | 1.243 | 1.625 |
| Radiation | 1.017 | 0.032 | 0.530 | 0.598 | 0.956 | 1.082 |

Other covariates also played a critical role in the CSS. Older age (45-64yrs) was associated with increased mortality (HR = 1.44, 95% CI: 1.00–2.07), while being married was protective (HR = 0.65, 95% CI: 0.58–0.74). Racial disparities were evident, with Non-Hispanic Black people having a higher risk of mortality (HR = 1.10, 95% CI: 1.00–1.22) compared to Non-Hispanic White people. Socioeconomic factors, such as living in large metropolitan areas, were associated with improved survival (HR = 0.74, 95% CI: 0.63–0.87). Advanced cancer stages were strongly associated with worse outcomes, with stage IV having the highest mortality risk (HR = 26.99, 95% CI: 11.85–61.46). Additionally, treatment factors such as receiving a prostatectomy significantly reduced the risk of mortality (HR = 0.38, 95% CI: 0.33–0.43), while chemotherapy and radiation were associated with increased mortality risk (HR = 1.62, 95% CI: 1.39–1.89; HR = 1.20, 95% CI: 1.09–1.33, respectively).

### Factors associated with Overall survival

In table 3, it reports the factors associated with the overall survival among individuals with prostate cancers in both states and in both time periods. The post-2015 period was associated with improved overall survival, with a hazard ratio (HR) of 0.90 (95% CI: 0.82–0.99), indicating a significant reduction in the risk of overall mortality during the period. The interaction between state and post-Medicaid expansion period showed that individuals in Kentucky had a reduction in mortality post-ACA (HR = 0.87, 95% CI: 0.75–1.01), although this effect was marginally significant (p = 0.065).

**Table 3:**
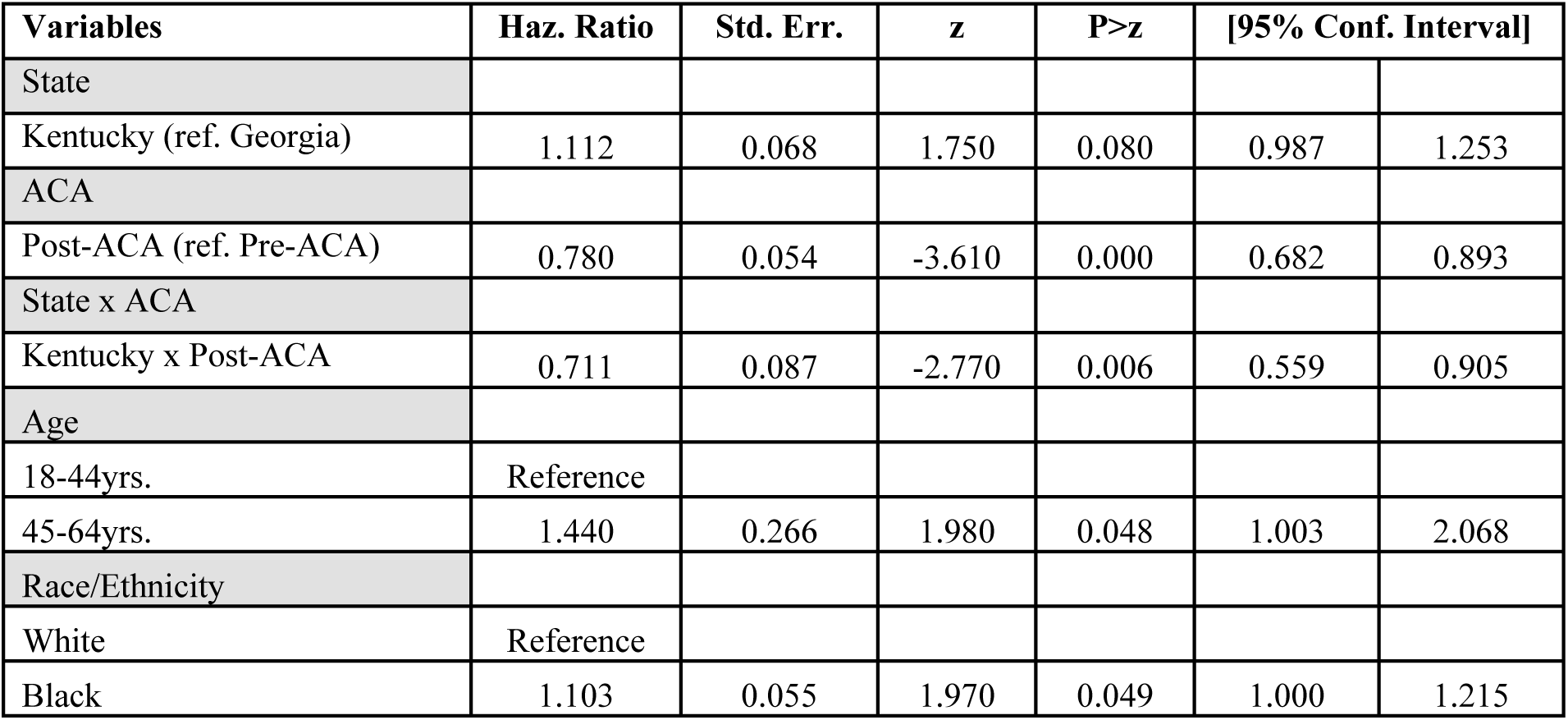
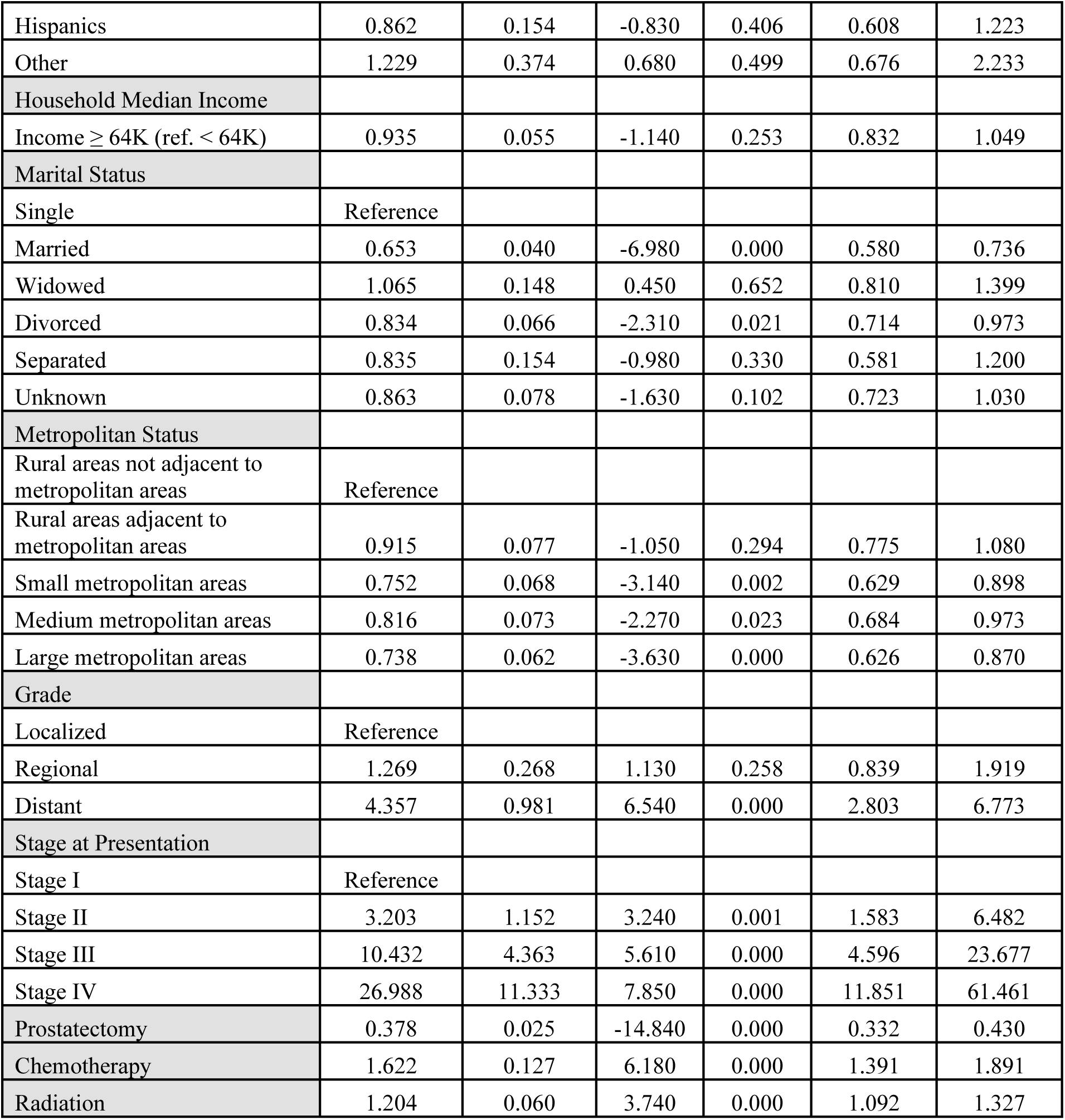
Cox Proportional Hazards Model Assessing Factors Associated with Overall Survival among Individuals with Prostate Cancer.

| Variables | Haz. Ratio | Std. Err. | z | P>z | [95% Conf. Interval] |  |
| --- | --- | --- | --- | --- | --- | --- |
| State |  |  |  |  |  |  |
| Kentucky (ref. Georgia) | 1.112 | 0.068 | 1.750 | 0.080 | 0.987 | 1.253 |
| ACA |  |  |  |  |  |  |
| Post-ACA (ref. Pre-ACA) | 0.780 | 0.054 | -3.610 | 0.000 | 0.682 | 0.893 |
| State x ACA |  |  |  |  |  |  |
| Kentucky x Post-ACA | 0.711 | 0.087 | -2.770 | 0.006 | 0.559 | 0.905 |
| Age |  |  |  |  |  |  |
| 18-44yrs. | Reference |  |  |  |  |  |
| 45-64yrs. | 1.440 | 0.266 | 1.980 | 0.048 | 1.003 | 2.068 |
| Race/Ethnicity |  |  |  |  |  |  |
| White | Reference |  |  |  |  |  |
| Black | 1.103 | 0.055 | 1.970 | 0.049 | 1.000 | 1.215 |
| Hispanics | 0.862 | 0.154 | -0.830 | 0.406 | 0.608 | 1.223 |
| Other | 1.229 | 0.374 | 0.680 | 0.499 | 0.676 | 2.233 |
| Household Median Income |  |  |  |  |  |  |
| Income ≥ 64K (ref. < 64K) | 0.935 | 0.055 | -1.140 | 0.253 | 0.832 | 1.049 |
| Marital Status |  |  |  |  |  |  |
| Single | Reference |  |  |  |  |  |
| Married | 0.653 | 0.040 | -6.980 | 0.000 | 0.580 | 0.736 |
| Widowed | 1.065 | 0.148 | 0.450 | 0.652 | 0.810 | 1.399 |
| Divorced | 0.834 | 0.066 | -2.310 | 0.021 | 0.714 | 0.973 |
| Separated | 0.835 | 0.154 | -0.980 | 0.330 | 0.581 | 1.200 |
| Unknown | 0.863 | 0.078 | -1.630 | 0.102 | 0.723 | 1.030 |
| Metropolitan Status |  |  |  |  |  |  |
| Rural areas not adjacent to metropolitan areas | Reference |  |  |  |  |  |
| Rural areas adjacent to metropolitan areas | 0.915 | 0.077 | -1.050 | 0.294 | 0.775 | 1.080 |
| Small metropolitan areas | 0.752 | 0.068 | -3.140 | 0.002 | 0.629 | 0.898 |
| Medium metropolitan areas | 0.816 | 0.073 | -2.270 | 0.023 | 0.684 | 0.973 |
| Large metropolitan areas | 0.738 | 0.062 | -3.630 | 0.000 | 0.626 | 0.870 |
| Grade |  |  |  |  |  |  |
| Localized | Reference |  |  |  |  |  |
| Regional | 1.269 | 0.268 | 1.130 | 0.258 | 0.839 | 1.919 |
| Distant | 4.357 | 0.981 | 6.540 | 0.000 | 2.803 | 6.773 |
| Stage at Presentation |  |  |  |  |  |  |
| Stage I | Reference |  |  |  |  |  |
| Stage II | 3.203 | 1.152 | 3.240 | 0.001 | 1.583 | 6.482 |
| Stage III | 10.432 | 4.363 | 5.610 | 0.000 | 4.596 | 23.677 |
| Stage IV | 26.988 | 11.333 | 7.850 | 0.000 | 11.851 | 61.461 |
| Prostatectomy | 0.378 | 0.025 | -14.840 | 0.000 | 0.332 | 0.430 |
| Chemotherapy | 1.622 | 0.127 | 6.180 | 0.000 | 1.391 | 1.891 |
| Radiation | 1.204 | 0.060 | 3.740 | 0.000 | 1.092 | 1.327 |

Other covariates also played a crucial role in overall survival. Age between 45-64yrs. was significantly associated with higher mortality (HR = 2.17, 95% CI: 1.69–2.81), while being married was also protective (HR = 0.65, 95% CI: 0.61–0.70). Racial disparities were evident, with Non-Hispanic Black people having a higher risk of mortality (HR = 1.10, 95% CI: 1.04– 1.17) compared to Non-Hispanic White people. Socioeconomic factors, such as living in large metropolitan areas, were associated with improved survival (HR = 0.71, 95% CI: 0.65–0.78). Advanced cancer stages were strongly associated with worse outcomes, with stage IV having the highest mortality risk (HR = 4.05, 95% CI: 2.99–5.48). Additionally, treatment factors such as receiving a prostatectomy significantly reduced the risk of mortality (HR = 0.47, 95% CI: 0.44– 0.51), while chemotherapy was associated with increased mortality risk (HR = 1.42, 95% CI: 1.24–1.62).

### Predicted Probabilities (CSS)

Table 4 reveals the predicted probabilities of the hazards of death for CSS between Georgia and Kentucky in the pre-ACA and post-ACA across different races/ethnicities and in the overall populations. In both states and across all races and ethnicities, the post-2015 period was associated with an improvement in CSS. Specifically, for the overall population, the probability of death decreased from 5.41 (95% CI: 1.55-9.28) in the pre-ACA period to 4.22 (95% CI: 1.10-7.35) in the post-ACA period in Georgia and from 6.02 (95% CI: 1.63-10.41) to 3.34 (95% CI: 0.80-5.88) in Kentucky. Among Non-Hispanic White people, the probabilities followed a similar trend, with the post-ACA period showing reduced risks compared to the pre-ACA period, such as in Kentucky, where the probability decreased from 5.79 (95% CI: 1.58-10.00) to 3.21 (95% CI: 0.78-5.65). Non-Hispanic Black people also experienced a reduction in the predicted probabilities of death post-ACA, with a decrease from 6.39 (95% CI: 1.69-11.09) to 3.54 (95% CI: 0.82-6.27) in Kentucky. Hispanic people, Non-Hispanic Asian/Pacific Islander people, and Native American people also showed a reduction in the post-ACA period across both states.

**Table 4:** Predicted Probabilities of Cancer-Specific Survival Among Individuals Prostate Cancer.

| Variables | ALL | White | Black | Hispanic | NHAPI | Native Americans |
| --- | --- | --- | --- | --- | --- | --- |
| Georgia x Post-ACA | 5.41 (1.55-9.28) | 5.21 (1.49-8.93) | 5.74 (1.61-9.88) | 4.49 (0.94-8.04) | 6.40 (0.44-12.36) | 2.42 (-2.63-7.47) |
| Georgia x Pre-ACA | 4.22 (1.10-7.35) | 4.07 (1.06-7.07) | 4.48 (1.15-7.82) | 3.51 (0.67-6.35) | 5.00 (0.25-9.75) | 1.89 (-2.06-5.84) |
| Kentucky x Post-ACA | 6.02 (1.63-10.41) | 5.79 (1.58-10.00) | 6.39 (1.69-11.09) | 5.00 (0.98-9.01) | 7.12 (0.41-13.83) | 2.69 (-2.94-9.32) |
| Kentucky x Pre-ACA | 3.34 (0.80-5.88) | 3.21 (0.78-5.65) | 3.54 (0.82-6.27) | 2.77 (0.47-5.08) | 3.95 (0.13-7.77) | 1.49 (-1.64-4.63) |

### Predicted Probabilities (Overall Survival)

Table 5 shows the predicted probabilities of overall survival between Georgia and Kentucky in the pre-ACA and post-ACA periods across racial and ethnic groups. In the overall population, the post-2015 period in Georgia experienced a reduction in the probability of death from 1.06 (95% CI: 0.81-1.31) pre-ACA to 0.95 (95% CI: 0.70-1.21) post-ACA. Similarly, in Kentucky, the probability of death decreased from 1.25 (95% CI: 0.94-1.56) pre-ACA to 0.98 (95% CI: 0.70-1.26) post-ACA. Among Non-Hispanic White people, a similar trend was observed, with the pre-ACA probability decreasing from 1.02 (95% CI: 0.78-1.26) to 0.92 (95% CI: 0.67-1.17) in Georgia and from 1.21 (95% CI: 0.91-1.51) to 0.94 (95% CI: 0.67-1.22) in Kentucky.

**Table 5:**
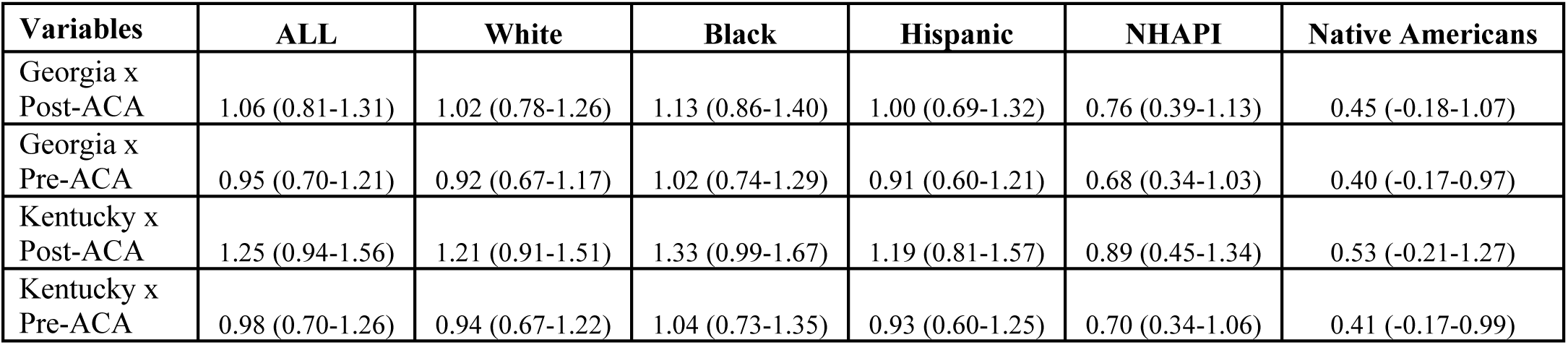
Predicted Probabilities of Cancer-Specific Survival Among Individuals Prostate Cancer.

For Non-Hispanic Black people, the predicted probability of death post-ACA decreased from 1.13 (95% CI: 0.86-1.40) to 1.02 (95% CI: 0.74-1.29) in Georgia and from 1.33 (95% CI: 0.99-1.67) to 1.04 (95% CI: 0.73-1.35) in Kentucky. Hispanic people also experienced a decrease in the probability of death, particularly in Kentucky, where the post-ACA period saw a reduction from 1.00 (95% CI: 0.69-1.32) to 0.91 (95% CI: 0.60-1.21). This pattern was also seen among Non-Hispanic Asian/Pacific Islander people and Native Americans people.

### DID for Cancer-Specific Survival

Table 6 reports the DID output on the change in the hazard of death for CSS among individuals with prostate cancer. Overall, the coefficient for all individuals was −1.49 (95% CI: −3.00 to 0.02, p=0.053), indicating a marginally significant reduction in the hazard of death. When broken down by race/ethnicity, Non-Hispanic White people showed a similar reduction (Coef. = −1.44, 95% CI: −2.89 to 0.01, p=0.052), as did Non-Hispanic Black people (Coef. = −1.58, 95% CI: −3.19 to 0.02, p=0.054), both nearing statistical significances. Hispanic people also experienced a reduction in hazard (Coef. = −1.24, 95% CI: −2.55 to 0.08, p=0.065), though the result was slightly less significant. The reduction was more pronounced for NHAPI people (Coef. = −1.76, 95% CI: −3.84 to 0.31, p=0.096), though it did not reach statistical significance. Native Americans showed the least reduction in hazard, with a coefficient of −0.67 (95% CI: −2.14 to 0.81, p=0.376), indicating no significant change.

**Table 6:** DID Output for Cancer-Specific Survival by Race/Ethnicity.

| Race/Ethnicity | Std. Err. | z | P>z | [95% Conf. Interval] |  |
| --- | --- | --- | --- | --- | --- |
| All | -1.492 | 0.770 | 0.053 | -3.000 | 0.017 |
| White Person | -1.436 | 0.740 | 0.052 | -2.885 | 0.014 |
| Blacks Person | -1.583 | 0.820 | 0.054 | -3.191 | 0.025 |
| Hispanic Person | -1.238 | 0.671 | 0.065 | -2.554 | 0.078 |
| Asian & Pacific Islanders | -1.764 | 1.059 | 0.096 | -3.839 | 0.311 |
| Native Americans | -0.667 | 0.753 | 0.376 | -2.142 | 0.808 |

### DID for Overall Cancer Survival

Table 7 reveals the DID results on the change in the hazard of death for overall survival among individuals with prostate cancers. There was a substantive reduction in mortality risk across most racial and ethnic groups post-Medicaid expansion in Kentucky compared to Georgia. For the overall population, the analysis showed a modest but statistically significant decrease in the hazard of death, with a coefficient of −0.17 (95% CI: −0.33 to −0.01, p=0.039). This trend was consistent across Non-Hispanic White people (Coef. = −0.16, 95% CI: −0.32 to −0.01, p=0.039) and Non-Hispanic Black people (Coef. = −0.18, 95% CI: −0.35 to −0.01, p=0.039) people, both of which experienced significant reductions in mortality risk. Hispanic people also experienced a significant reduction in hazard (Coef. = −0.16, 95% CI: −0.31 to −0.01, p=0.043). Non-Hispanic Asian/Pacific Islander people showed a similar, though marginally significant, reduction (Coef. = −0.12, 95% CI: −0.24 to 0.00, p=0.060). Native Americans had the smallest reduction, with a coefficient of −0.07 (95% CI: −0.19 to 0.05, p=0.245), indicating no significant change in overall survival.

**Table 7:** DID Output for Overall Survival by Race/Ethnicity.

| Race/Ethnicity | Coefficient | Std. Err | P>z | [95% Conf. Interval] |  |
| --- | --- | --- | --- | --- | --- |
| All | -0.168 | 0.081 | 0.039 | -0.326 | -0.009 |
| White Person | -0.162 | 0.078 | 0.039 | -0.315 | -0.008 |
| Black Person | -0.179 | 0.086 | 0.039 | -0.348 | -0.009 |
| Hispanic Person | -0.159 | 0.079 | 0.043 | -0.313 | -0.005 |
| Asian & Pacific Islanders | -0.120 | 0.064 | 0.060 | -0.245 | 0.005 |
| Native Americans | -0.071 | 0.061 | 0.245 | -0.190 | 0.048 |

## DISCUSSION

Overall, there was a decline in the predicted probabilities of cancer-specific deaths in the post-2015 period, observed across both states regardless of Medicaid expansion status. However, this trend was not mirrored in the risk of overall deaths, as no significant difference was noted between the pre- and post-Medicaid expansion periods for overall survival in either state. While a more pronounced reduction in cancer-specific mortality was noted in Kentucky compared to Georgia, the difference was not statistically significant. Conversely, there was a significant 16.8 percentage point decrease in overall mortality risk among individuals with prostate cancer in Kentucky relative to Georgia, with the reduction ranging from −32.6% to −9%. This improvement in overall survival was consistent across different racial/ethnic groups and was particularly notable among Black individuals, who experienced a 17.9 percentage point drop in the risk of overall deaths compared to their counterparts in Georgia. Other significant predictors of improved cancer-specific and overall survival included younger age, White race, higher household median income, married status, and residence in metropolitan areas. Undergoing prostatectomy was significantly associated with improved survival, while radiation therapy did not show a significant effect. In contrast, chemotherapy was linked to worse cancer-specific and overall survival, potentially due to higher disease burden among individuals receiving this treatment modality.

The improvement in survival among individuals diagnosed with prostate cancer in recent years is likely multifactorial. Advances in early detection and screening methods, particularly widespread adoption of prostate-specific antigen (PSA) testing, have facilitated diagnosis at earlier, more treatable stages. Additionally, innovations in treatment modalities—including the development of more effective surgical techniques, radiotherapy, androgen deprivation therapy, and novel targeted therapies—have significantly enhanced disease management. Increased access to multidisciplinary care, improved supportive therapies, and heightened awareness of prostate cancer through public health campaigns have also contributed to better outcomes. Furthermore, there has been an overall improvement in healthcare delivery systems and a greater emphasis on personalized treatment approaches tailored to patients’ genetic and molecular profiles, which may account for the observed survival gains.

When comparing the change in prostate cancer-specific mortality between Kentucky and Georgia, there were no statistically significant differences, although the results indicated a trend towards a greater decline in Kentucky. This suggests that while there may be a modest reduction in prostate cancer-specific deaths following the implementation of the ACA and Medicaid expansion in Kentucky, the findings do not provide robust evidence of a substantive improvement in outcomes relative to Georgia.

The absence of a substantive impact of Medicaid expansion on prostate cancer outcomes in Kentucky compared to Georgia may be attributed to several factors. Prostate cancer is generally a slow-growing malignancy with a favorable prognosis, and the benefits of improved healthcare access may take years to manifest in survival outcomes. The indolent nature of the disease, coupled with high baseline survival rates, could dilute the measurable effects of policy changes within the study timeframe. Moreover, while we observed a trend towards a greater decline in prostate cancer-specific mortality in Kentucky, the lack of statistical significance suggests that the improvement may be influenced by other concurrent healthcare advancements or socioeconomic factors not directly related to Medicaid expansion. These nuances indicate that longer follow-up periods or studies focusing on more aggressive cancers might be needed to detect the full impact of Medicaid expansion on cancer-specific outcomes.

Kentucky, a Medicaid expansion state, demonstrated a significant 16.8% reduction in overall mortality risk compared to Georgia, a non-expansion state. This is particularly notable because overall mortality, unlike cancer-specific mortality, may more comprehensively capture the broader benefits of Medicaid expansion, such as improved access to preventive care, management of comorbidities, and timely interventions for acute conditions. Given the generally indolent nature of prostate cancer, the impact of Medicaid expansion on prostate cancer-specific deaths may take longer to manifest, whereas the significant reduction in all-cause mortality in Kentucky suggests that enhanced healthcare access had an immediate and measurable effect on broader health outcomes.

This finding is important in light of the fact that no statistically significant change in overall mortality was observed across the study period in either state, emphasizing that the mortality reduction in Kentucky may be directly attributable to increased healthcare access following the implementation of the ACA and its Medicaid expansion component. These results highlight the potential of Medicaid expansion to reduce health disparities and improve survival, particularly in populations that were previously uninsured or underinsured, reinforcing the role of policy-driven healthcare reforms in enhancing population health outcomes.

Interestingly, individuals who received chemotherapy experienced a significantly higher hazard of both cancer-specific and overall mortality. This may be explained by a higher disease burden and more advanced cancer stage at the time of treatment, as chemotherapy is often reserved for high-risk or metastatic cases. Additionally, the observed mortality risk may reflect an imbalance in sociodemographic and clinical factors, as patients receiving chemotherapy were more likely to belong to vulnerable groups such as low-income families, unmarried or single individuals, and racial minorities—populations that often have limited access to comprehensive healthcare and may present with more aggressive disease. Moreover, disparities in treatment adherence and supportive care in these groups could further contribute to poorer outcomes, underscoring the need for tailored interventions to address these inequities and improve survival among chemotherapy-treated patients.

This study highlights persistent disparities in cancer outcomes, with Black individuals, those from the lowest-income households, single and unmarried individuals, and residents of rural areas facing the highest risk of both cancer-specific and overall mortality. These disparities may be driven by a combination of factors, including delayed diagnosis, limited access to high-quality care, and a greater burden of comorbidities in these populations. Additionally, socioeconomic barriers and geographical challenges often result in reduced access to advanced treatments and follow-up care, further exacerbating survival disparities.

### STRENGHT AND LIMITATIONS

This study has several strengths, including the use of the SEER registry, a high-quality, population-based cancer database that provides comprehensive clinical and demographic information, allowing for robust analysis of cancer outcomes. A key strength is our approach of comparing two states with similar demographic profiles—Kentucky and Georgia—rather than grouping states broadly into Medicaid expansion versus non-expansion categories. This approach mitigates the potential bias introduced by heterogeneity in Medicaid implementation timelines and healthcare infrastructure across states, enhancing the specificity of our findings.

Furthermore, the use of a DID approach allows for stronger causal inferences from observational data by accounting for underlying time trends, thus providing a more accurate estimate of the impact of Medicaid expansion on survival outcomes. However, our study has limitations, including the inability to control pre-existing comorbidities or baseline insurance status, which may influence both treatment decisions and survival outcomes. Additionally, unmeasured factors such as differences in healthcare delivery systems, patient adherence, and provider practices could also affect mortality and potentially confound our results. Despite these limitations, our study offers valuable insights into the nuanced effects of Medicaid expansion on cancer outcomes and overall mortality.

In conclusion, while we did not observe a significant difference in prostate cancer-specific mortality or survival between Kentucky and Georgia, Kentucky’s Medicaid expansion was associated with a notable improvement in overall survival, evidenced by a significant reduction in all-cause mortality compared to Georgia. This finding underscores the broader impact of Medicaid expansion on health outcomes beyond cancer-specific measures, highlighting the policy’s potential to reduce overall mortality through improved healthcare access. However, significant disparities persist, with Black individuals, low-income households, rural residents, and unmarried individuals continuing to experience poorer outcomes in both cancer-specific and overall survival. These results emphasize the need for targeted strategies to address these inequities and ensure that the benefits of expanded healthcare coverage reach the most vulnerable populations.

## Data Availability

The data supporting the findings of this study are available on request from the corresponding author.

